# Sentinel Event Surveillance to Identify Threats to SMC Effectiveness: Association Between Breakthrough Infections, SMC Uptake, Adherence and Parasite Drug Resistance

**DOI:** 10.64898/2026.11.10.26367777

**Authors:** Dancan M. Wakoli, Beatrice Rotich, Edwin Mbugua, Suzanne Van Hulle, Joseph Kipkoech, Lucy Abel, Thomas Katairo, Emmah Kimachas, Amy Wesolowski, Melissa Conrad, Diana Menya, Wendy Prudhomme O’Meara

**Affiliations:** Academic Model Providing Access to Healthcare, Eldoret, Kenya; Catholic Relief Services; Duke Global Health Institute, Durham, NC, USA; Infectious Disease Research Center, Kampala, Uganda; Johns Hopkins University Bloomberg School of Public Health, Baltimore MD USA; Moi University College of Health Sciences, Eldoret, Kenya; Duke University School of Medicine, Durham, NC, USA

## Abstract

Seasonal malaria chemoprevention (SMC) is an important malaria prevention strategy used widely in areas of West Africa with highly seasonal transmission. Monthly administration of sulfadoxine-pyrimethamine plus amodiaquine (SPAQ) to young children during the high-transmission season substantially reduces malaria morbidity and mortality. SMC has recently been introduced in northern Kenya, where pastoralist communities experience intense malaria transmission. However, implementation in this setting presents unique challenges, including population mobility, vast distances, dispersed settlements, and high levels of parasite resistance to SP in parts of the region. Effective surveillance is needed to identify gaps in SMC delivery and uptake and to monitor threats to program effectiveness.

We implemented an innovative sentinel-site surveillance approach in which every malaria case among SMC-eligible children at selected health facilities was investigated to identify gaps in SMC delivery, uptake, and effectiveness. We also assessed the prevalence of parasite alleles associated with SP and AQ resistance and examined their relationship with SMC uptake.

Household travel was substantially more common than caregiver refusal among reported reasons for missing an SMC cycle. More than one-third of children presenting with malaria had missed their most recent SMC cycle, while 15% were within a few days of receiving the next scheduled cycle. The prevalence of SP resistance mutations was considerably lower than reported in other parts of East Africa: only 13.4% of infections carried the quintuple mutant haplotype IRNI/GEAA (PfDHFR 51I, 59R, 108N, PfDHPS 437G, 540E) associated with high level resistance to SP. However, the prevalence of resistance-associated mutations was higher among children who had recently received SMC. The allele associated with reduced susceptibility to AQ was not detected.

Sentinel surveillance identified geographic and implementation gaps in real time, allowing program managers to target SMC distribution and community sensitization. Although the prevalence of resistance-associated mutations that could threaten SPAQ effectiveness was low, evidence of intrahost selection among children who had received SMC suggests that continued molecular surveillance will be important as SMC is scaled up in northern Kenya. This sentinel surveillance approach provides a practical framework for monitoring SMC implementation in remote, highly mobile populations while simultaneously tracking emerging threats to SPAQ effectiveness.

## Introduction

Seasonal malaria chemoprevention has been expanded substantially in recent years, reaching 54 million children across 20 countries in 2024 (1). Evidence from both randomized controlled trials and observational studies in real world programmatic implementation demonstrate that SMC is beneficial in preventing morbidity and severe disease in young children, with coverage and effectiveness varying across settings (2–7).

Although SMC has yielded meaningful progress, several factors undermine the effectiveness of SMC. Children under 5 continue to account for a substantial burden even in countries where SMC has been implemented at scale for more than a decade. SMC programs are compromised by the logistical challenge of reaching every eligible child (8) during peak transmission periods, and those reached are required to complete a 3 day course. Limited adherence to the 3-day regimen (8,9) resulting in suboptimal drug and consequentially increased risk of selection of drug resistant parasites (10) which may in turn lead to decreased protective effectiveness.

SMC was implemented in Turkana, Northern Kenya for the first time in 2024. Despite the semi-nomadic lifestyle of the target populations, it reached 97% of eligible children with at least one cycle and 71% with all five cycles (11), predominantly through door to door distribution by community health promoters (CHPs) supplemented with fixed points, mobile fixed points and integrated health outreaches in more remote sparsely populated villages. Ultimately, malaria risk was reduced by over two thirds.

Robust surveillance systems are lacking in high or moderate malaria transmission regions of Sub-Saharan Africa, which bear the greatest burden of malaria. This study focused on children presenting with a positive malaria diagnosis during the 2025 transmission season to identify underlying reasons for suboptimal protection during SMC. The overall goal was to use surveillance to investigate reasons for illness and improve the SMC program in real time.

## Materials and Methods

### Study area and sentinel facilities

Turkana County is the largest county in Kenya but is sparsely populated and predominantly rural. Turkana residents are primarily nomadic pastoralists whose movement is shaped by the arid environment and the seasonal rainfall. Despite the arid climate and low population density, malaria transmission is very high, exceeding 400 cases per 1000 people in an average season. In 2024, Turkana County piloted SMC in one subcounty to protect young children from malaria infection. After the success of the first campaign, Turkana County implemented its second round of SMC from June-October 2025. As in the previous round, sulfadoxine-pyrimethamine and Amodiaquine (SPAQ) was delivered over a five-day interval to all children between 3-59 months in Turkana Central subcounty. The first cycle was initiated on 05/06/2025 and repeated every 28 days for five total cycles.

Sentinel surveillance was initiated on 10/07/2025, immediately following the second cycle of SMC. Enrollment ended on 28/10/2025, 28 days after the final day of the final cycle of SMC. Six public health facilities that covered a wide geographical area including a peri-urban area within the main town, communities near Lake Turkana, and those serving populations residing in very rural areas which offer outreach services to remote communities, were selected for this study.

### Participant recruitment

A sentinel event was defined as an SMC-eligible child aged 3-59 months who lived in the SMC implementing sub county (Turkana Central) between July-October 2025 and had a positive malaria diagnosis by malaria rapid diagnostic test (mRDT) or microscopy at one of the participating facilities. Children presenting to the facility who were referred for a malaria diagnostic test were screened for eligibility. The accompanying parent or guardian was invited to participate in the study if the child met the criteria for enrollment. A trained nurse or clinical officer sought written informed consent from the parent or guardian.

### Data and sample collection

A standardized questionnaire was administered in KOBOCollect to capture basic information including age, geographical residence, bed net use, history of malaria-like illness, and SMC information. SMC questions explored whether SMC cycles were missed and why, the date of the last SMC drugs, adherence to the 3-day regimen, adverse reactions and vomiting during drug administration. Data were entered directly into tablets and synced to an encrypted password-protected server. For children older than 6 months, a Dried Blood Spot (DBS) was prepared by dropping 5-10 µL of blood on to a filter paper and allowing it to dry completely. Children under 6 months of age were exempted from DBS sample collection, but their caregivers participated in the questionnaire. The study protocol was reviewed by the Moi Teaching and Referral Hospital/Moi University Institutional Scientific and Ethics Review Committee (Approval number FAN:0005076).

### *Plasmodium falciparum* detection and sequencing

Genomic DNA (gDNA) was extracted from dried blood spot (DBS) samples using the Chelex 100 method as previously described, with minor modifications (12,13). Briefly, a single DBS punch was placed into a 96-deep-well plate and treated with saponin and proteinase K before DNA extraction with Chelex 100. *Plasmodium falciparum* was detected using a TaqMan real-time PCR assay targeting specific alleles of the *Pfdhfr* and *Pfdhps* genes on a QuantStudio 3 Real-Time PCR System (Applied Biosystems, Foster City, CA, USA). Each 12 µL reaction contained 6 µL Bio-Rad SsoAdvanced™ Universal Probes Supermix, 0.15 µL each of forward and reverse primers, 0.18 µL probe, 4.52 µL nuclease-free water, and 1 µL gDNA template. Five standards (1, 2, 10, 100, and 1000 parasites/µL), prepared as described (13), and nuclease-free water were included in duplicate as positive and negative controls, respectively. Samples with cycle threshold (Ct) value <40 were considered positive for *P. falciparum* (14). Parasite densities were estimated from a standard curve generated using the standards (13).

Libraries were prepared from all *P. falciparum*-positive samples using the paragon primer pools M1.1 and M2.1 from the Multiplex Amplicons for Drug, Diagnostic, Diversity, and Differentiation Haplotypes using Targeted Resequencing (MAD^4^HatTeR) protocol (15,16). Sequencing was performed on pooled indexed libraries using the Illumina MiSeq platform to assess for sulfadoxine-pyrimethamine and amodiaquine (SPAQ) resistance markers at the Infectious Diseases Research Centre (IDRC), Central Public Health Laboratory, Uganda, as previously described (16). Sequences have been submitted to NCBI Sequence Read Archive (project number pending).

### Statistical analysis

Data were cleaned in STATA 17 (StataCorp LLC, College Station, TX, USA) and visualized in R version 4.6.1. Participant characteristics were summarized, and SMC uptake ascertained at the conclusion of each SMC cycle. Participants were categorized according to SMC uptake, children who received all cycles since the onset of surveillance, those who had missed some cycles and those who had not received SMC at all. Reasons for sub-optimal SMC either due to missed cycles or inadequate drug concentration were ascertained. Resident villages of sentinel cases were shared with program implementers to enable real time correction of problems with drug delivery or logistical challenges affecting reach. Children with confirmed malaria illness by PCR were included in subsequent analysis to investigate probable causes of malaria during SMC. Sequencing data were analyzed using the MAD^4^HatTeR bioinformatics pipeline (version 0.2.1) (15), to determine genotypes at loci associated with susceptibility to components of SPAQ. Sequencing depth and per-sample coverage were calculated as described (16). Specifically, loci with a total coverage of at least 100 reads were considered successfully genotyped; within these loci, minor alleles were included for analysis if they had a within-sample allele frequency (WSAF) > 0.75% and were supported by at least 10 reads. Genotypes were then classified based on these filtered allele frequencies as wild type, mutant, or mixed. Lastly, the prevalence of each resistance SNP and haplotype was calculated as a proportion of successfully genotyped samples using Stata version 17 (StataCorp LLC, College Station, TX, USA).

## Results

### Study population

Between 10 July – 28 October 2025, 707 SMC-eligible children aged 3-59 months were enrolled. 181 participants who were reported positive by microscopy had a negative PCR result and were subsequently excluded from data analysis. Given the high rate of false positives by microscopy, a small number of samples that could not be tested by PCR but were positive by microscopy were also excluded (n=18).

Gender and age of enrolled children was similar across the surveillance sites (Table 1), but a distinct difference was observed in location of the sleeping space, with children from rural communities (Kalakol, Kapua, Nadoto and Namakuse) sleeping both indoors and outdoors. Overall, 92% of children had a mosquito net available for their sleeping space. Of those who had a net, 93% (n=437/469) reported sleeping under the net the night before.

**Table 1.** Demographic characteristics of children testing positive for malaria at surveillance sites.

|  | Kalakol GOK | Kanamkemer SC Hosp | Kapua Dispensary | Nadoto Dispensary | Ngiitakito Dispensary | Namakuse Dispensary | Total |
|---|---|---|---|---|---|---|---|
| N | 101 (19.9%) | 96 (18.9%) | 21 (4.1%) | 133 (26.2%) | 98 (19.3%) | 59 (11.6%) | 508 (100.0%) |
| **Age (months)** |  |  |  |  |  |  |  |
| 3-5 | 2 (2.0%) | 1 (1.0%) | 0 (0.0%) | 1 (0.8%) | 2 (2.0%) | 0 (0.0%) | 6 (1.2%) |
| 6-11 | 11 (10.9%) | 9 (9.4%) | 2 (9.5%) | 8 (6.0%) | 4 (4.1%) | 1 (1.7%) | 35 (6.9%) |
| 12-59 | 85 (84.2%) | 85 (88.5%) | 17 (81.0%) | 121 (91.0%) | 92 (93.9%) | 58 (98.3%) | 458 (90.2%) |
| Above 59 | 3 (3.0%) | 1 (1.0%) | 2 (9.5%) | 3 (2.3%) | 0 (0.0%) | 0 (0.0%) | 9 (1.8%) |
| **Gender** |  |  |  |  |  |  |  |
| Female | 44 (43.6%) | 45 (46.9%) | 9 (42.9%) | 76 (57.1%) | 46 (46.9%) | 28 (47.5%) | 248 (48.8%) |
| Male | 57 (56.4%) | 51 (53.1%) | 12 (57.1%) | 57 (42.9%) | 52 (53.1%) | 31 (52.5%) | 260 (51.2%) |
| **Any SMC?** |  |  |  |  |  |  |  |
| No | 9 (8.9%) | 14 (14.6%) | 0 (0.0%) | 1 (0.8%) | 12 (12.2%) | 7 (11.9%) | 43 (8.5%) |
| Yes | 92 (91.1%) | 82 (85.4%) | 21 (100.0%) | 132 (99.2%) | 86 (87.8%) | 52 (88.1%) | 465 (91.5%) |
| **Sleeping location** |  |  |  |  |  |  |  |
| Indoors | 56 (55.4%) | 86 (89.6%) | 3 (14.3%) | 105 (78.9%) | 96 (98.0%) | 13 (22.0%) | 359 (70.7%) |
| Outdoors | 2 (2.0%) | 2 (2.1%) | 0 (0.0%) | 28 (21.1%) | 1 (1.0%) | 1 (1.7%) | 34 (6.7%) |
| Both | 43 (42.6%) | 8 (8.3%) | 18 (85.7%) | 0 (0.0%) | 1 (1.0%) | 45 (76.3%) | 115 (22.6%) |
| **Have a net?** |  |  |  |  |  |  |  |
| No | 14 (13.9%) | 7 (7.3%) | 0 (0.0%) | 1 (0.8%) | 13 (13.3%) | 4 (6.8%) | 39 (7.7%) |
| Yes | 87 (86.1%) | 89 (92.7%) | 21 (100.0%) | 132 (99.2%) | 85 (86.7%) | 55 (93.2%) | 469 (92.3%) |
| **Slept under net last night** |  |  |  |  |  |  |  |
| No | 29 (28.7%) | 13 (13.5%) | 0 (0.0%) | 3 (2.3%) | 15 (15.3%) | 11 (18.6%) | 71 (14.0%) |
| Yes | 72 (71.3%) | 83 (86.5%) | 21 (100.0%) | 130 (97.7%) | 83 (84.7%) | 48 (81.4%) | 437 (86.0%) |

### SMC uptake and adherence

The number of SMC cycles received was captured for each participant who presented over the transmission season. Over 90% of participants received at least 1 cycle of SMC (Table 1) and 48% received all cycles for which they were eligible (Supplemental Table 1). Few caregivers reported children receiving zero cycles of SMC (n=43, 8.5%). The reasons reported for any missed SMC cycles (N=578) included that the household was not visited by a drug distributor (n=96, 16.6%) or the fixed point distribution was too far away (23, 4%), the caretaker or the child travelled during the SMC distribution days (218, 37.7%) or that the caregivers chose for their child not to participate in that cycle (n=79, 13.7%). Some children missed a cycle because they were not eligible on the day of distribution due to illness (139, 24%).

On day 1, children receive SP and AQ. Drug distributors demonstrate to caregivers how to administer the remaining two doses of AQ on day 2 and day 3. Reported adherence to day 2 and 3 doses among enrolled children was high. Of those who received at least one cycle of SMC, 94.6% of caregivers reported giving AQ on day 2 and day 3 as required while 3.5% gave only one of the 2 remaining doses and 1.9% of caregivers did not give any doses after day 1.

We further explored the relationship between the date of the last SMC dose and the date when they were diagnosed with malaria (Fig 1). Thirty-two percent (31.6%) of children who had at least one cycle of SMC had their last dose of SMC more than 32 days earlier, which equates to missing their last dose of SMC. An additional 17.3% were enrolled 26-31 days post-SMC, during the period when they were awaiting the next cycle. It is possible that many of these were referred to the health facility by the drug distributor if they were found unwell during the campaign. About 12% of participants were enrolled within 7 days of their last reported SMC dose. Some of these children may have had a non-malaria illness (or even SPAQ side effects) but also harbored an asymptomatic infection that was still detectable by mRDT when they reported to the health facility.

**Fig 1.**
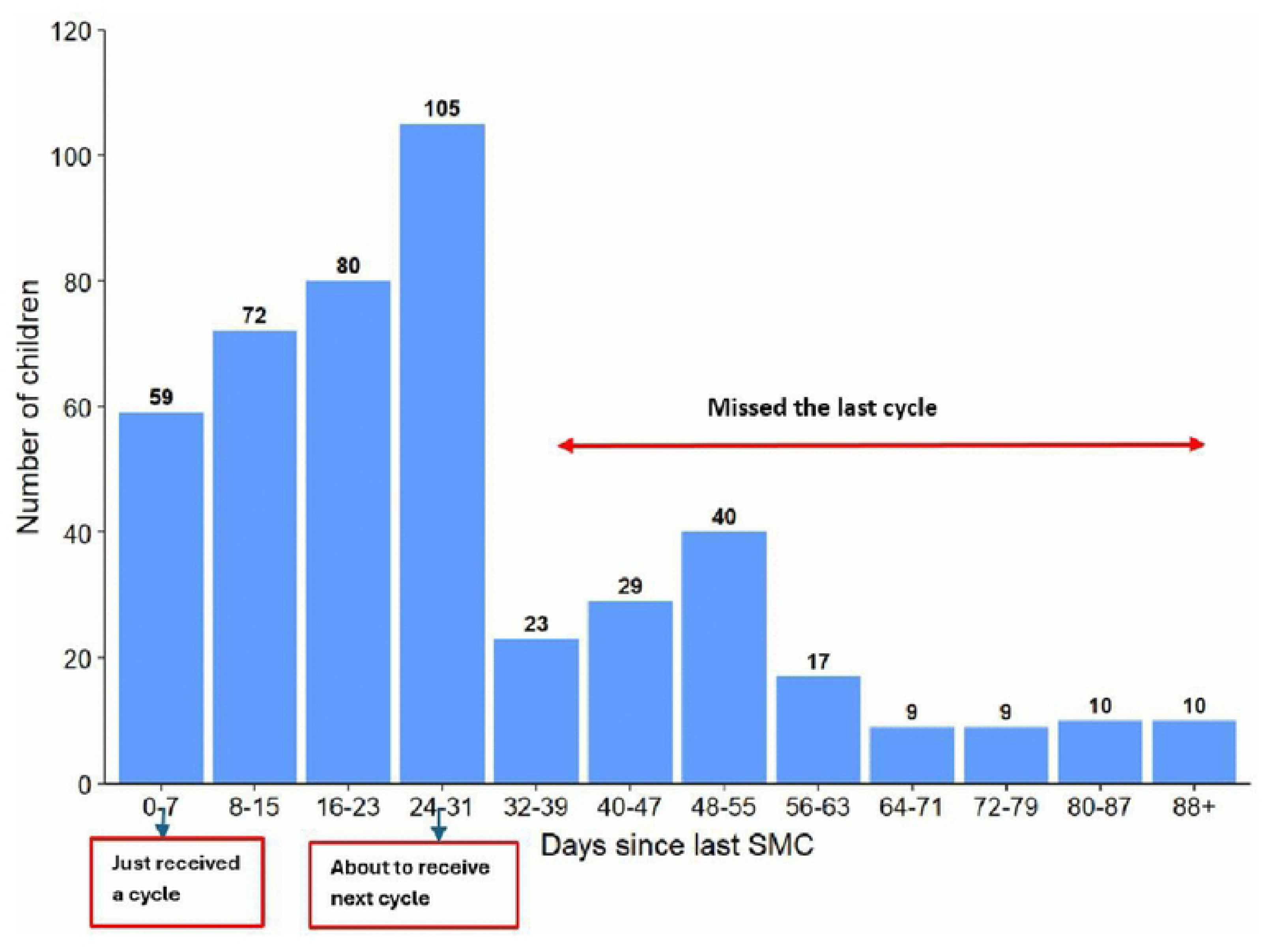
Relationship between malaria diagnosis and times since the last SMC dose.

### Possible reasons for breakthrough infection

A substantial proportion of children who tested positive for malaria either had no cycles of SMC in 2025 (8.7%) or had missed the last cycle of SMC (SMC > 32 days ago, 24.7% (Fig 2). They therefore remained vulnerable to malaria infection. An additional 9.8% likely had inadequate SPAQ on board for protection, either due to non-adherence to day 2/3 doses of AQ by the caregiver (4.9%) or they vomited their first dose of drugs within 30-40 minutes of ingestion with no repeat dose (4.9%). These children may not have had drug levels within the therapeutic range to eliminate existing infections or prevent new ones. Overall, 14.8% of children were enrolled 26-31 days post-SMC, potentially a window of waning protection as they awaited the next cycle or identified as febrile at home and referred to the facility by the drug distributor. Over 40% (41.9%) of enrolled children fell ill during the window of peak protection with no other explanations for diminished protection based on their reported SMC uptake.

**Fig 2.**
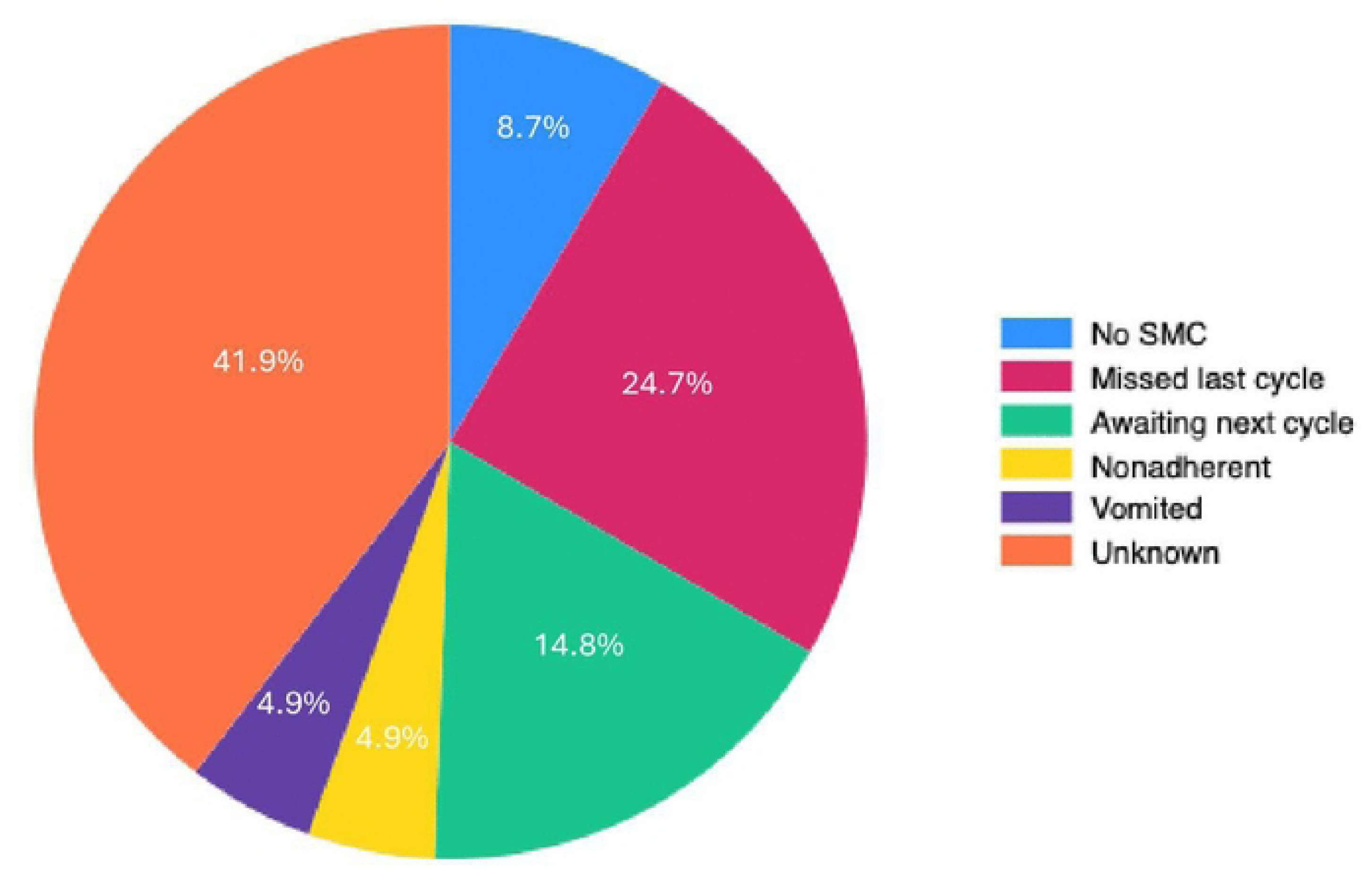
Distribution of possible reasons for having a malaria infection during the SMC period. Each child has only one most likely reason assigned.

### Parasite resistance to sulfadoxine-pyrimethamine plus amodiaquine

448 samples were further analyzed for known genetic markers of parasite resistance to SP and AQ, of which 369 were successfully genotyped (Table 2). The prevalence of initial mutations required for pyrimethamine resistance but not associated with treatment failure were at fixation (98.9% PfDHFR 51I, 99.2% PfDHFR 108N). More than half of infections were fully wildtype at PfDHFR 59 (51.7%) and 100% of infections had the wildtype allele at PfDHFR 164. Resistance mutations in PfDHPS were less prevalent; 437G was present in 29.4% of samples and 540E in only 23.3%. PfDHPS 581 and 613 were fully wildtype. The quintuple mutant haplotype IRNI/GEAA (PfDHFR 51I, 59R, 108N, PfDHPS 437G, 540E) associated with moderate levels of treatment failure were present in only 13.4% of infections (Table 3). The sextuple mutant haplotype (PfDHFR 51I, 59R, 108N and PfDHPS 437G, 540E, 581G) associated with high level resistance, was completely absent. The PfDHFR mutation 164L which is associated with high level treatment failure particularly in East Africa was also absent.

**Table 2.** Frequency of drug resistance haplotypes among children enrolled in surveillance. *If all sequenced alleles were wild type, then wildtype was assumed if the sequence was missing for PfDHFR 164 and PfDHPS 581. If PfDHFR 108 was missing, mutant was assumed. For all samples with mixed alleles, mutant type was called in the variant.

| **Variant** | **Prevalence (%)** |
|---|---|
| **PfDHFR** |  |
| N51I (n=356) |  |
| Wild-type | 4 (1.1%) |
| Mutant | 352 (98.9%) |
| C59R (n=356) |  |
| Wild-type | 184 (51.7%) |
| Mixed allele | 45 (12.6%) |
| Mutant | 127 (35.7%) |
| S108N (n=357) |  |
| Wild-type | 3 (0.8%) |
| Mutant | 354 (99.2%) |
| I164L (n=357) |  |
| Wild-type | 357 (100.0%) |
| **PfDHPS** |  |
| I431V (n=385) |  |
| Wild-type | 385 (100%) |
| A437G (n=369) |  |
| Wild-type | 261 (70.7%) |
| Mixed allele | 32 (8.7%) |
| Mutant | 76 (20.6%) |
| K540E (n=369) |  |
| Wild-type | 283 (76.7%) |
| Mixed allele | 27 (7.3%) |
| Mutant | 59 (16.0%) |
| A581G (n=369) |  |
| Wild-type | 369 (100.0%) |
| A613S (n=355) |  |
| Wild-type | 354 (99.7%) |
| Mutant | 1 (0.3%) |
| **PfMDR** |  |
| N86Y |  |
| Wild-type | 365 (100%) |
| Y184F |  |
| Wild-type | 152 (40.1%) |
| Mixed | 43 (11.2%) |
| Mutant | 184 (48.5%) |
| D1246Y |  |
| Wild-type | 379 (100%) |
| **PfCRT** |  |
| M74I (n=368) |  |
| Wild-type | 353 (95.9%) |
| Mixed | 7 (1.9%) |
| Mutant | 8 (2.1%) |
| N75E (n=368) |  |
| Wild-type | 353 (95.9%) |
| Mixed | 7 (1.9%) |
| Mutant | 8 (2.1%) |
| K76T (n=368) |  |
| Wild-type | 353 (95.9%) |
| Mixed | 7 (1.9%) |
| Mutant | 8 (2.1%) |

**Table 3.** Prevalence of DHFR and DHPS resistance marker haplotypes.

| **Haplotypes\*** | **Prevalence (%)** |
|---|---|
| PfDHFR variants (n=351) |  |
| NCSI | 3 (0.9%) |
| ICSI | 0 (0%) |
| ICNI | 178 (50.7%) |
| IRNI | 170 (48.4%) |
| IRNL | 0 (0%) |
| PfDHPS variants (n=369) |  |
| AKAA | 261 (70.7%) |
| GKAA | 22 (6.0%) |
| GEAA | 86 (23.3%) |
| Quintuple mutant (n=351) |  |
| IRNI/GEAA | 47 (13.4%) |

No mutations were found in *P. falciparum* multi-drug resistance gene 1 (PfMDR1) position 86 or 1246 (Table 2). PfMDR1 Y184 was present in 51.5% (195/379) of infections. The PfCRT CVIET haplotype (C72, V73, M74I, N75E, K76T), which was common in Africa before artemether-lumefantrine replaced chloroquine as the first-line therapy, was seen in 15 of 368 samples. Ten of these were collected from children 26 or more days after their last SMC cycle and all had SP resistance mutations. Of these, 9 were quintuple mutants in PfDHFR and PfDHPS (GEAA/IRNI), while an additional 5 had the GEAA/ICNI genotype.

The prevalence of resistance markers varied by time from last SPAQ dose (Fig 3). PfDHPS 437G increased from 14.3% in children who had never received SMC and gradually increased up to 34.0% in children who received SMC 26-31 days prior (p=0.04). Similarly, the PfDHPS 540E allele was more common in children who had received SMC than those who had not (p=0.02). Other alleles did not vary from time since last SMC. The prevalence of PfDHPS 437G, 540E and PfDHFR 59R were slightly higher in children who had vomited their last dose of SPAQ compared to those who did not receive SMC, but the numbers were too small to establish statistically significant differences (Fig 4).

**Fig 3.**
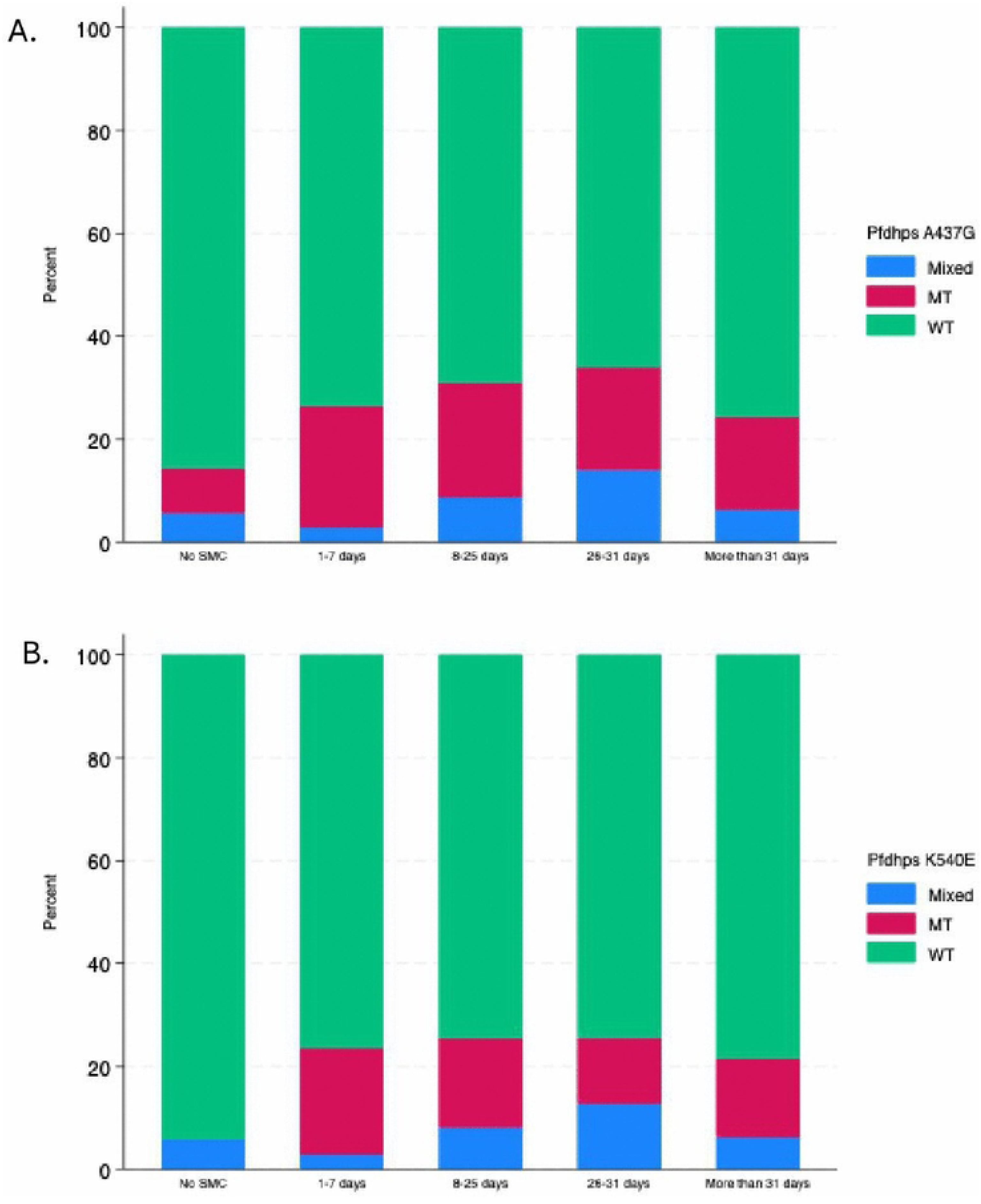
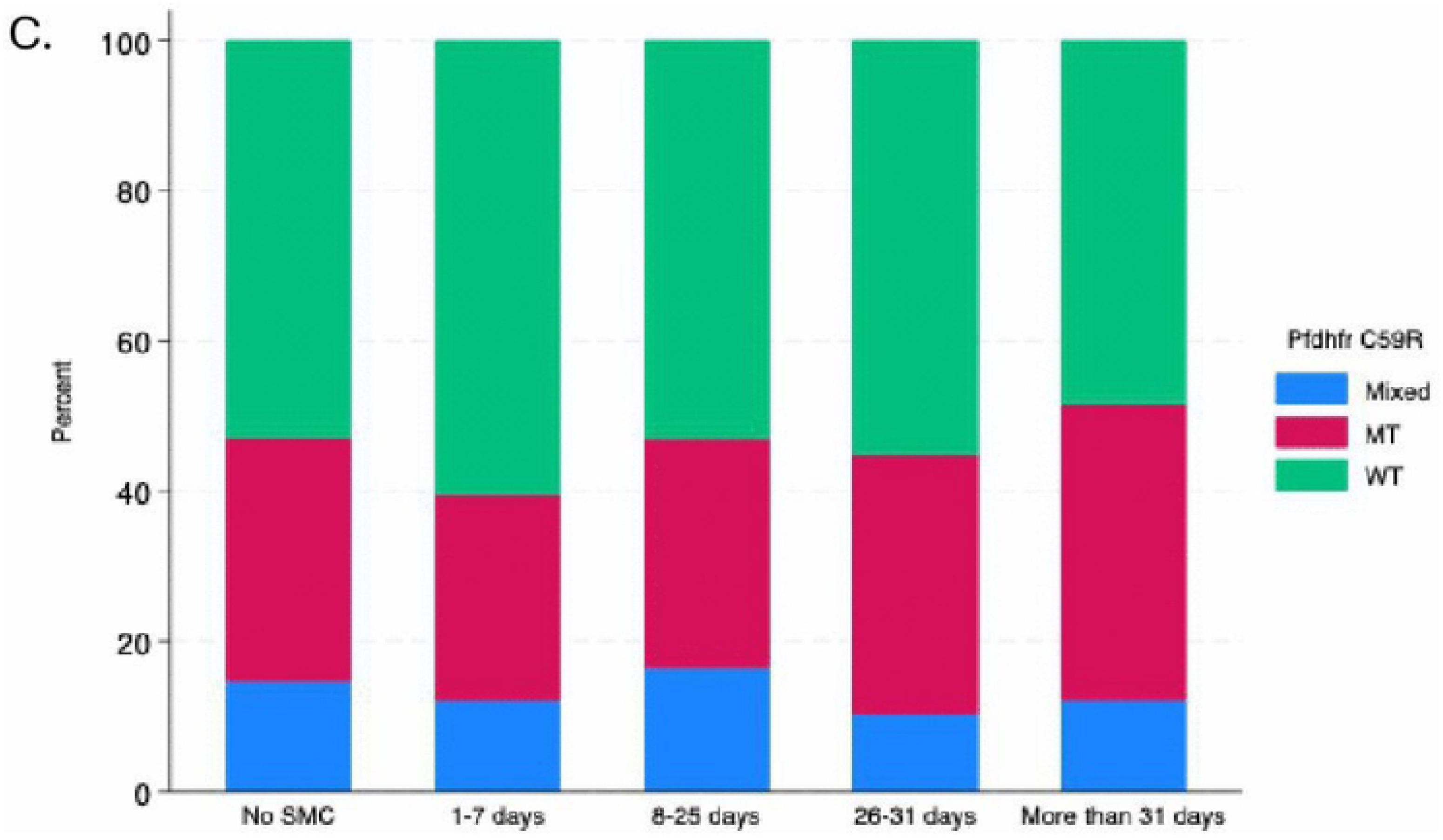
Percentage of infections with Mutant, Wildtype or Mixed alleles by time since last dose of SMC. (A) PfDHPS A437G (B) PDHPS K540F and (C) PFDHFR C59R.

**Fig 4.**
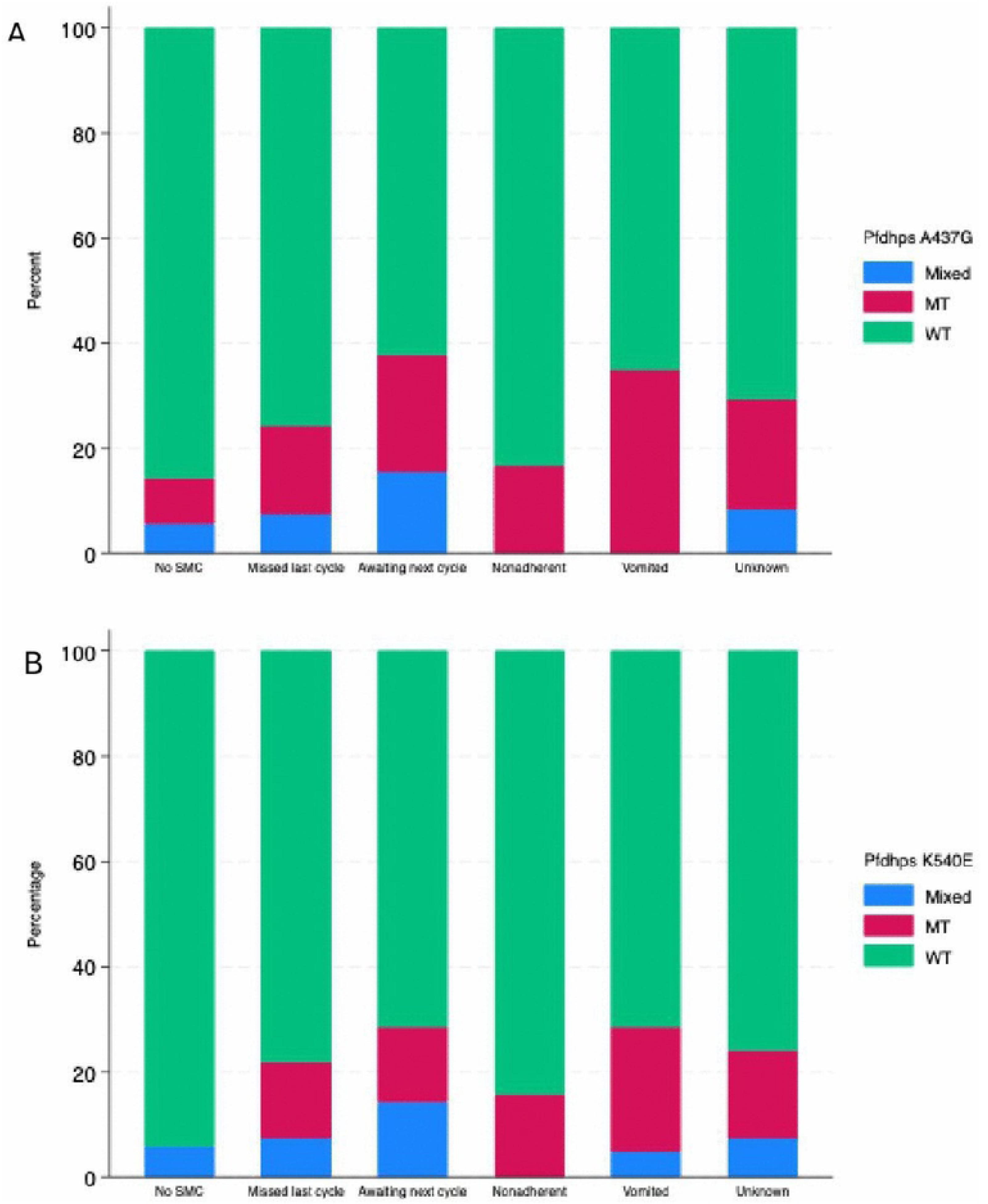
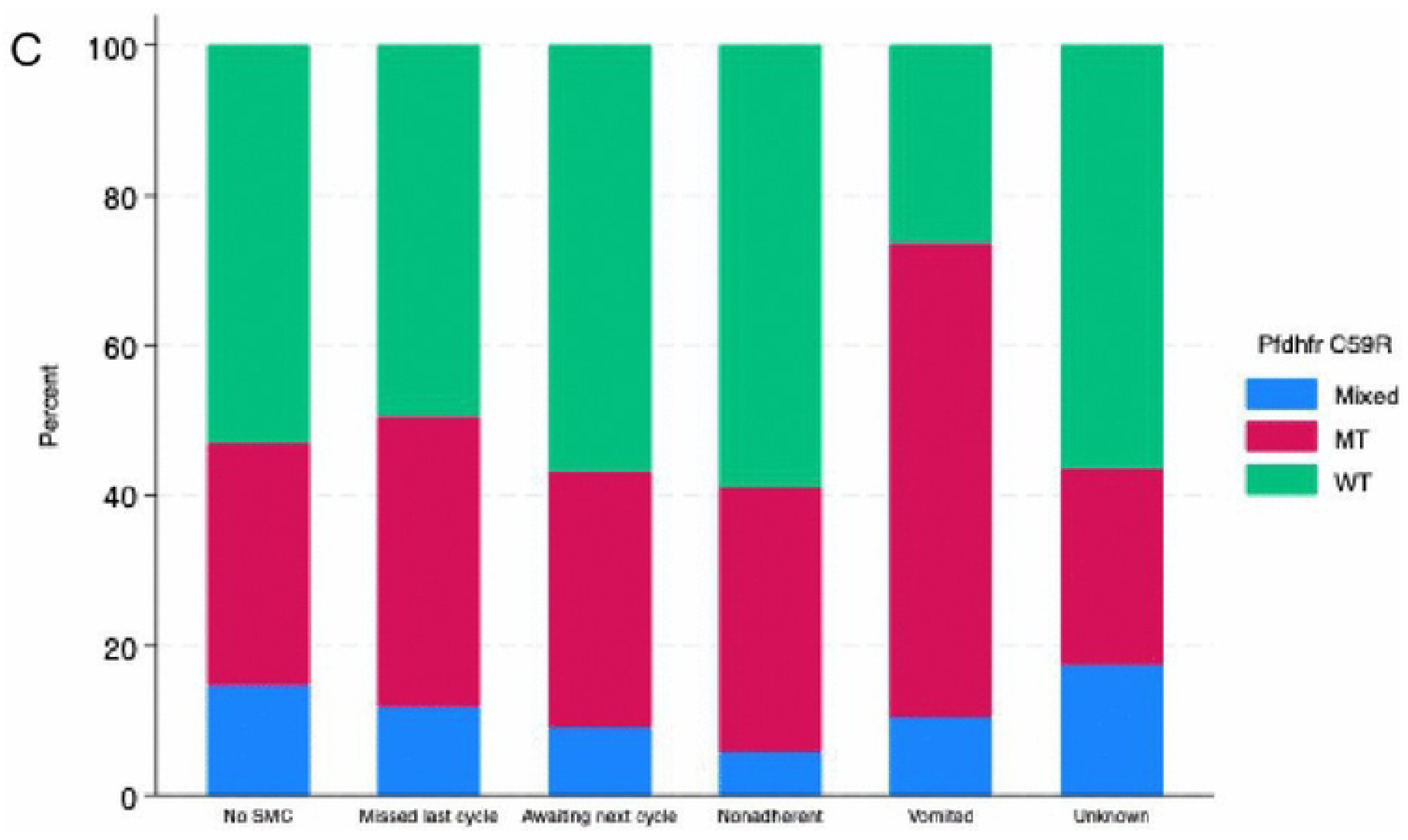
Percentage of infections with Mutant, Wildtype or Mixed alleles by possible reason for diminished protection. (A) PfDHPS A437G (B) PfDHPS K540E and (C) PfDHFR C59R.

## Discussion

Our report of facility-based surveillance demonstrates the value of using case investigation at sentinel sites to inform SMC delivery. By identifying children with confirmed malaria infection during the SMC campaign, we were able to identify geographic areas with low coverage and document the challenge that very remote communities and vast distances pose for SMC in this context. A fifth of children who missed a cycle were affected by inability of the drug distributor to reach them or the long distance to the nearest fixed point. Frequent travel by these highly mobile communities was cited as the main reason for nearly 40% of missed SMC cycles, much higher than reported refusal rates.

SMC guidelines require that any child experiencing malaria-like symptoms on the day of SMC delivery should not be given SMC unless they test negative for malaria by either RDT or microscopy (17–19). When community-based testing is unavailable, as in Turkana, all children who are unwell are referred to the facility and not given SMC at home. Often those referrals are not completed and consequently that child will not receive SMC. The finding that nearly a quarter of missed SMC cycles was due to illness at the time of distribution suggests two priority areas for improving SMC coverage in subsequent campaigns. First, aligning the first cycle of SMC to begin before significant seasonal transmission could reduce the number of children who were already infected and therefore ineligible for SMC. Second, strengthening community-based malaria testing could facilitate delivery of SMC to malaria-negative children experiencing symptoms on the day of distribution. Several countries have successfully integrated testing and treatment by CHPs into their SMC campaigns (18,20,21).

A missed cycle of SMC accounted for a large proportion of breakthrough infections. This was in line with earlier findings from a multi-country study in west Africa, where between 15-89% of infected children had missed their most recent SMC cycle (Cairns et al., 2020). Similarly, another study found that 82% of children presenting with malaria shortly after a previous SMC cycle had no detectable SP drug, implying that missing cycles contributes substantially to breakthrough infections (10). In our study, there was a substantial proportion of children for whom a specific reason such as non-adherence or missed cycles could not be identified. We investigated parasite sensitivity to SMC drugs as a possible reason for intercurrent infections.

Overall, the prevalence of genetic markers associated with resistance to SP were moderate, but the most highly resistant parasites commonly found in the western part of Kenya and other parts of East Africa were absent (23–27). Resistance-associated mutations in *PfDHPS* at position 437 and 540 (24,28) were found in fewer than 30% of infections, but the proportion was higher in children who had received SMC, potentially indicating within-host selection. A similar pattern was reported for an SMC cohort in Turkana where parasites in breakthrough infections were enriched with resistance markers in children who received SMC compared to those who did not (29). In contrast, SMC eligible children with malaria in Burkina Faso had higher prevalence of several SP-resistance alleles, including *PfDHPS* 613S, but prevalence was not associated with recent SMC or SP drug levels in the blood (10). In our study, the proportion of parasites harboring *PfDHPS* 437G was highest in infections nearing the end of the SP protective window, possibly due to the declining drug concentrations over time to a level permissible for parasites with low-level drug tolerance. This is consistent with recent findings showing that the duration of SP protection is dependent on the *PfDHPS* genotype, with substantially shorter protection against parasites carrying 437G and additional *dhps* mutations compared with SP susceptible parasites (30). Children who had vomited their SMC dose had the highest proportion of resistant infections, but the total number of children in this category was small (n=20 vomited with sequenced infections) and the difference was not statistically significant.

Another possible explanation for infections in children receiving SMC is inadequate drug absorption leading to suboptimal levels in the blood and shorter protective windows. Several studies have investigated SP and AQ blood levels in SMC-eligible children and found lower than expected drug levels in some children even when the research team directly observed administration of all three doses (31,32). In one study, SP and AQ levels generally agreed with parent self-report of adherence, but were significantly lower in those who did not receive directly observed therapy for all three doses, indicating some over-reporting by caretakers (32). SP and AQ blood levels were not associated with moderate malnutrition (10), the prevalence of which is high in Turkana children and has been shown to alter the pharmacokinetics of SP absorption (33).

A small number of infections harbored mutations in PfCRT (the CVIET variant) associated with diminished sensitivity to amodiaquine, albeit at a lower prevalence than other areas of Kenya (34) and much lower than observed in pooled analysis of west African isolates following two years of SMC (35). Most of these were identified in children who had SMC more than three weeks prior and are potentially selected for by subtherapeutic concentrations of amodiaquine. However, the changes in PfCRT (74I, 75E and 76T) alone may not be adequate to alter sensitivity to amodiaquine in the absence of additional mutations in PfMDR (86Y, 1246Y) which were not present in any infections (34,36–39). The observation As has been documented in other parasite populations in East Africa, roughly half of infections harbored a mutation in the multi-drug resistant gene PfMDR at position 184 (34,40,41), resulting in the allele N86/F184/D1246 which is associated with high AL usage (34,42), although its effect on lumefantrine sensitivity versus parasite fitness is complex (43–45). AL is the first-line treatment for uncomplicated malaria in Kenya and is widely used. A similar distribution of PfMDR Y184F was observed in Burkina Faso where SMC has been used for nearly a decade (46) Importantly, the prevalence of the N86/F184/D1246 allele in our study was identical to previous studies from western Kenya where AL has been shown to remain highly effective (34,44).

This study investigated children who presented to health facilities with malaria in six sentinel facilities. We purposively selected facilities which serve different types of populations (pastoralist, peri-urban, fishing communities) and represent varying malaria burden. As a result, the findings may not be generalizable but rather indicative. We relied heavily on the caregiver report of uptake of SMC which may not be completely accurate, probably erring on the side of higher uptake and adherence. The finding that microscopy results included a large numbers of false positives is problematic generally, but specifically for SMC may lead to declining confidence in the efficacy of the treatment if children take SMC but are subsequently being diagnosed with and treated for malaria. In addition, treatment with AL between SMC cycles will often result in the child being ineligible for the subsequent SMC cycle due to AL consumption within the last two weeks. These children will be unprotected for weeks once the AL is cleared as they await the next cycle of SMC, further undermining the effectiveness of the SMC program.

Our results suggest that SMC with SPAQ is expected to be highly effective in this setting. Measures should be taken to preserve the useful lifespan of SPAQ and safeguard against increasing resistance by reinforcing the importance of adhering to the complete monthly regimen. Sentinel site monitoring through case investigation at health facilities can generate actionable information in real-time to assist SMC programming. It also provides a highly sensitive population for identifying changes in parasite genetic markers associated with drug sensitivity.

## Data Availability

The data are being uploaded onto NCBI SRA and will be publicly available

## Acknowledgement

We thank all study participants and caretakers for their contribution to this study, as well as the staff of the participating health facilities, and Turkana County Health Team, for their support. We also acknowledge Linda Maraga, Mark Amunga, and Millicent Cherono for their assistance with sample processing and laboratory testing. We thank Bienvenu Nsengimaana and Muwanika John Vester from IDRC, Uganda for their contributions to the sequencing work. Finally, we are grateful to Catholic Relief Services for support during this study.

## Supporting Information

S1 Table. Number and proportion of children missing cycles over the study period.

## Notes

### Competing Interest Statement

The authors declare that there are no competing interests

### Author Declarations

This study was approved by the Moi Teaching and Referral Hospital/Moi University Institutional Scientific and Ethics Review Committee (Approval number FAN:005076).

